# Seroprevalence and risk factors of brucellosis in pastoralists and their livestock in South Sudan

**DOI:** 10.1101/2024.04.10.24305653

**Authors:** Emmanuel P. Lita, Erneo B. Ochi, Gerald Misinzo, Henriette van Heerden, Robab Katani, Jacques Godfroid, Coletha Mathew

**Author notes:** Corresponding author Emmanuel P. Lita.

## Abstract

**Background:** Brucellosis poses serious public health implications and substantial economic losses in pastoral rural settings in South Sudan. In humans, brucellosis is almost always originating from animals. Current literature provides scant data regarding the seroprevalence of brucellosis in South Sudan. This cross-sectional study investigates the seroprevalence of brucellosis among the pastoral community and livestock and identifies risk factors for the disease from two counties, Terekeka and Juba in Central Equatoria State (CES), South Sudan.

**Methodology:** A total of 986 serum samples from humans (n=143), cattle (n=478), sheep (n=86), and goats (n=279) were randomly collected from 17 livestock camps in CES. Serum samples for the humans, cattle and goats were screened for *Brucella-*specific antibodies using rose Bengal plate test (RBPT) and further confirmed by competitive enzyme linked immune sorbent assay (c-ELISA) in series. All the sera from sheep were tested in parallel using RBPT and c-ELISA. A camp was considered positive when at least one animal of either species tested positive on the c-ELISA. Univariate analysis using binary logistic regression with a confidence interval of 95% at a p-value of ≤ 0.05 was used to identify the association between the potential individual risk factors and *Brucella* seropositivity. The investigated risk factors for livestock included age, sex, species, prior abortion history, retained placenta, parity, and reproductive status. Variables found to have associations in univariate analysis (*p* =0.25) with *Brucella* seropositivity were further included in multivariable logistic regression. The risk factors investigated for humans included, gender, age, educational level, occupation, marital status, drinking of raw milk, aiding female animals during delivery, eating of undercooked meat and blowing of air into the cow’s uterus through the vagina.

**Results:** The study revealed seroprevalence of 21.7%, 11.8%, and 4.8% in cattle, goats, and humans, respectively. Our results indicated that all sheep serum samples were negative on both RBPT and c-ELISA. The seroprevalence in the 13 camps from TerekekaCounty was 100% (13/13) compared to 50% (2/4) seropositive from 4 camps in Juba County. All the variables investigated in the univariate analysis of risk factors in cattle were significantly associated with *Brucella* seropositivity: sex (OR:4.5, 95% CI: 2.2 – 8.9, p:<0.001), age (OR:6.6, 95% CI: 2.3 –19.1, p:<0.001), abortion history(OR:3.1, 95% CI: 1.8 – 5.2, p:<0.001), retained placenta (OR:2.5, 95% CI: 1.4 – 4.4, p: 0.001),parity (OR:2.3, 95% CI: 1.1 – 4.7, p:0.020). However, in small ruminants, none of the potential risk factors were associated with *Brucella* seropositivity. In humans, blowing air through cow’s vagina (OR: 1.4, 95%CI: 0.782 – 2.434, p: 0.035) was the only variable found to be significantly associated with *Brucella* seropositivity at the univariate analysis. The forceful blowing of air into cow’s vagina to induce milk is a common practice among the pastoral communities in South Sudan.

The multivariable logistic regression model identified sex, age, and abortion history as statistically significant factors for *Brucella* seropositivity in cattle. The odds of seropositivity were nearly threefold (OR: 2.8; 95% CI: 1.3 – 5.8, p: 0.006) higher in cows compared to bulls (male cattle). Cattle over two years old had higher odds of *Brucella* seropositivity than young animals (OR: 3.5, 95% CI: 1.2 – 10.3-, p: 0.025). Cows with a history of abortion had higher odds of *Brucella* seropositivity (OR: 2.8, 95% CI: 1.6 – 4.7, p: <0.001).

**Conclusion:** This is the first study to report occurrence of brucellosis in goats and its absence in sheep in South Sudan. Altogether, our results suggest that *Brucella abortus* infecting primarily cattle has spilled over to goats but not (yet) to sheep. The present study also shows occurrence of brucellosis in cattle, goats and people in the pastoral community and recommends implementation of One Health approach for effective mitigation of this disease.

**Author summary:** Brucellosis is a neglected, bacterial zoonotic disease that is caused by several species of the genus Brucella. Cross-species transmission of Brucella can occur in mixed or integrated farming systems. The disease poses serious public health implications and substantial economic losses particularly in low-income countries including South Sudan.

This study was conducted to estimate the seroprevalence of brucellosis in pastoralists, their livestock as well identifying potential risk factors associated with *Brucella* infection. Knowledge of the seroprevalence of brucellosis and risk factors is a prerequisite towards planning an effective mitigation strategy of the disease.

The study revealed high seroprevalence of brucellosis in cattle compared to goats, and the following risk factors were identified; prior history of abortion, age (old) and sex (female) significantly associated with *Brucella* infection. Surprisingly, sheep were found to be seronegative.

## INTRODUCTION

Brucellosis is a significant zoonotic disease affecting many countries in sub-Saharan Africa including South Sudan. *Brucella* spp. are the etiological agents of the disease that affect both humans and animals[1]. The species of *Brucella* are well-adapted to their hosts, however, a spillover can occur when there is an intermingling of various species of animals[2] as in the case with the agro-pastoral and pastoral system practiced in South Sudan [3]. The disease affecting livestock and humans is caused by *B. melitensis* mainly in goats and sheep, *B. abortus* mainly in cattle and buffalo as well as *B. suis* in pigs [4].There are several predisposing factors attributed to the occurrence of brucellosis in humans and animals.

In livestock, the disease causes reduced milk production, longer calving intervals, abortions, still birth, swollen joints and infertility [5].

Transmission of brucellosis to humans occurs through the consumption of infected, unpasteurized animal-milk products, through direct contact with infected animal parts (such as the placenta by infection through bruised skin and mucous membranes), and inhalation of infected aerosolized particles [6]. In humans, clinical brucellosis presents as acute or sub-acute febrile illness and is characterized by intermittent fever accompanied by malaise, anorexia, and prostration[7].

The economic losses due to brucellosis are enormous and incur costs to humans either directly (e.g. health care costs for the diagnosis, treatment, and management of clinically ill patients) or indirectly (e.g. loss of work days, lost leisure time, loss of productive years due to premature death[8]. Studies have shown that brucellosis is high in pastoral and mixed farming system where humans have been embedded with livestock, so it constitutes a high risk of infection [9–13].

Furthermore, several studies within the African region have identified several risk factors and reported varying prevalence levels of brucellosis based on spatial and temporal features, diagnostic methods, and species. [9,10,13–15].

However, few studies have been conducted in South Sudan to assess the prevalence of brucellosis in humans and cattle[16–19].These studies have reported seroprevalence of 23.2% (48/250) [17]and 23.3% (97/416) [16]in cattle and humans, respectively. There were no reported studies on brucellosis in sheep and goats. Despite that, the pastoral communities usually keep their animals such as cattle, sheep, and goats, in the cattle camps. It is found that keeping different animal species plays a pivotal role in cross-species transmission and maintenance of brucellosis[20]; [21].There is inadequate knowledge of the epidemiology and risk factors of brucellosis in other livestock species and their role in transmission of the infection to humans in South Sudan.

Understanding these gaps in knowledge is a prerequisite for the development of effective mitigation measures for the disease in South Sudan.

Hence, this study estimates seroprevalence and identifies risk factors associated with *Brucella* sero-positivity among pastoralists and their cattle, sheep, and goats in Central Equatoria State (CES), South Sudan.

## MATERIALS AND METHODS

### Study area

The study was conducted in Terekeka and Juba Counties of CES, South Sudan. In Terekeka County, three Payams namely Reggo, Nyori and Terekeka were randomly selected for the study. A Payam is the second-lowest administrative division, below Counties, in South Sudan. Terekeka County is located on both east and west banks of the White Nile River north of Juba. The county includes low-lying swampy areas that usually flood but provide grazing in the dry season. Rainfall is about 900 millimeters annually. Livestock rearing is considered an important part of people’s livelihood in Terekeka County, CES. In Juba County, northern Bari and Munuki payams were randomly selected. Juba is the largest city of South Sudan located in the center of CES. It borders Terekeka County to the north and Kajo-keji and Lainya Counties to the South. Residents of Juba engage in diverse range of livelihood. A total of 17 cattle camps were randomly selected, of which 13 were from Terekeka County and four from Juba County as shown in Figure 1.

**Figure 1:**
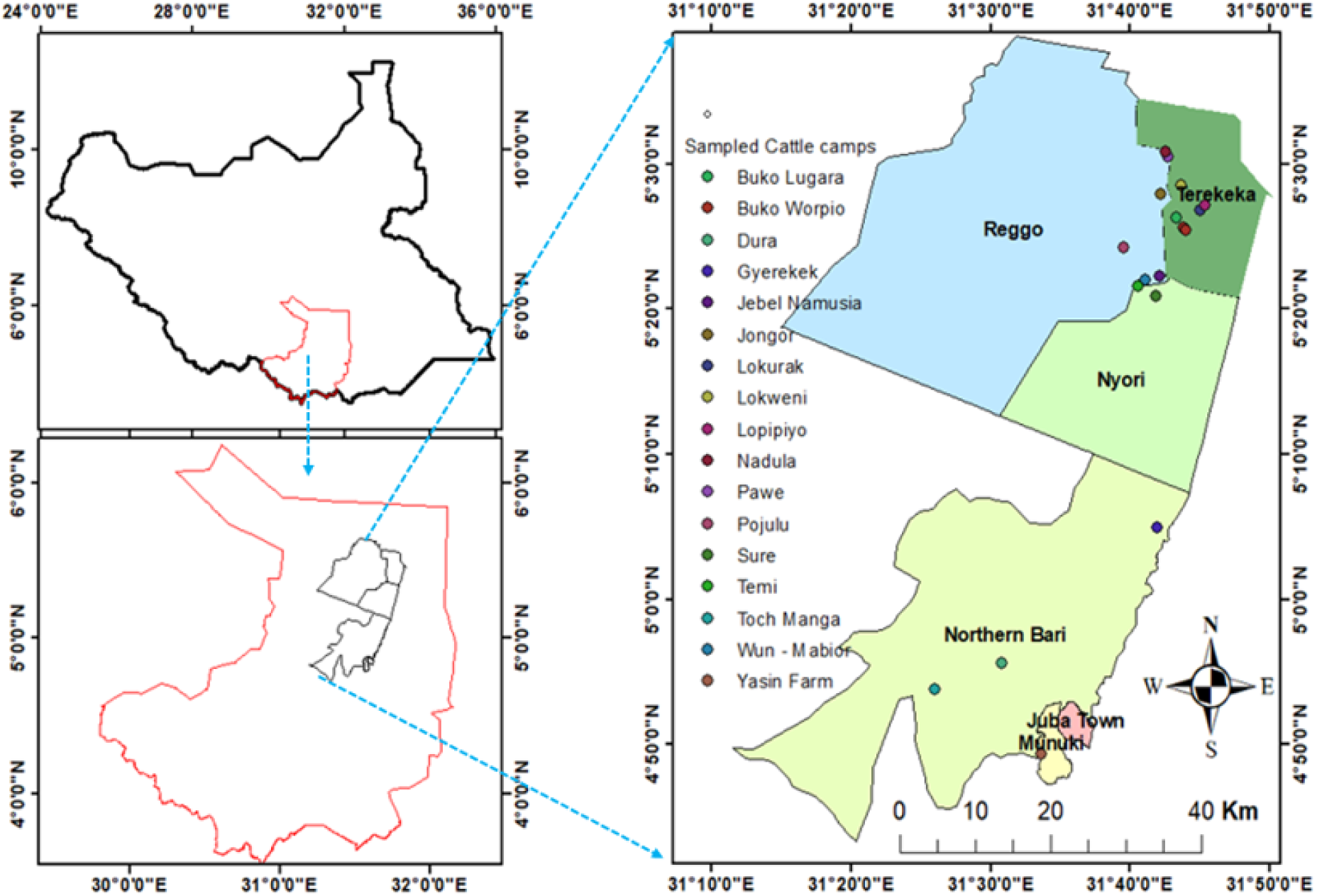
Location of the Central Equatorial State of South Sudan indicated in upper right with study area showing sampled cattle camps in Terekeka and Juba Counties in lower right consisting of Reggo, Terekeka Nyori in the Terekeka County and Northern Bari and Munuki Payams in the Juba County, indicated on left.

### Study design and subject

A cross-sectional design was planned using a multistage method of sampling for data collection. Briefly, cattle camps were identified and randomly selected; a proportional random sampling was then used to sample individuals. The study subjects comprised humans linked to cattle, ≥18 years old and of different genders. Livestock species, including cattle, sheep, and goats were sampled from ≥ 6 months old, and from different sexes. These animal species had no previous record of brucellosis vaccination and were mixed in the same cattle camps and managed entirely under a pastoral farming system.

### Sample size determination in animals and humans

The total sample required was determined according to formula given by[22].An expected individual animal prevalence (P) of 25.3% [19]and (P) of 50 % were used for calculating the sample size of cattle and small ruminants, respectively. The 50% expected prevalence for small ruminants was chosen because of no previous studies conducted to estimate prevalence of brucellosis in goats and sheep as per the context of South Sudan.

Based on the formula, 384 samples were to be collected from small ruminants and 290 from the cattle.

However, a total of 478 cattle, 86sheep and 279goats of different ages and sexes were included in this study.

In humans, a total of 143blood samples were collected from herders in the selected cattle camps who participated voluntarily.

### Blood collection and seroprevalence

A total of five to seven milliliters of blood were drawn aseptically from the jugular vein of each randomly selected animal using a needle and plain vacutainer tube. Immediately, the vacutainer tubes were labeled and coded and kept at room temperature overnight. The next day, the samples were centrifuged, and sera were harvested and placed into labeled cryovial tubes of 2 ml. A case history, detailed information about each animal sampled, and metadata were recorded in the data sheet.A medical technician collected a blood sample in humans, and sera separation followed the same protocol used in the animals. The collected sera were kept at -20 °C pending analysis.

### Rose Bengal Plate Test (RBPT)

The harvested sera from the humans, cattle, sheep and goats were all screened for *Brucella* antibodies using RBPT according to procedure described by[23]. The sera from the humans, cattle and goats were tested in series while the sheep sera were tested in parallel using RBPT and c-ELISA. The antigen was obtained from the Animal and Plant Health Agency (APHA), New Haw, Addlestone, Surrey, England.

A cattle camp was considered positive if at least one positive *brucella* reactor was found among the animals.

The test was conducted at the College of Veterinary Medicine and Biomedical Science, Sokoine University of Agriculture, Morogoro, Tanzania.

### Competitive Enzyme Linked Immuno Sorbent Assay (c-ELISA)

Sera found positive on the RBPT were further subjected on c-ELISA kit (Boehringer Ingelheim Svanonva, Uppsala, Sweden) for confirmation. The test was performed according to the protocol provided by the manufacturer with positive and negative controls. The samples were run in duplicates.

### Identification of Risk Factors of the disease

Univariate analysis and chi square test using a confidence interval of 95% at a p-value of ≤ 0.05 was used to identify the association between the potential individual risk factors and *Brucella* seropositivity. Risk factors associated with the disease were identified using multivariable logistic regression analysis of risk factors for *Brucella* seropositivity in cattle and small ruminants. Variables with a p-value≤ 0.25 from the univariate analysis were included in the multivariable analysis. Backward stepwise (Wald) model was used and the validity of the test was assessed by computing Hosmer-Lemeshow goodness-of-fit.

### Data management and statistical analysis

The study used both qualitative and quantitative methods of data collection and analysis. Data was analyzed using Statistical Package for Social Sciences (SPSS) version 20. Descriptive statistic wasrun to obtain the frequency distribution and percentages and univariate analysis was computed to identify association between variables.

## RESULTS

### Socio-demographic characteristics of studied pastoralist

A total of 143 pastoralist comprising females 9.0% (13/143) and males 91% (130/143) were included in the study. The majorities of the participants were single 68% (97/143) and have not attended formal education84% (120/145).The age category “18 – 25 years old” 63% (90/143) was the majority, followed by age group ˃32 years old 22.3% (32/143).

### Overall seroprevalence of brucellosis in different animal species

A total of 143 human sera and 843 of livestock comprising of cattle (478), sheep (86) and goats (279), of different age and sexes were screened for anti-*Brucella* antibodies. The seroprevalence in humans revealed 4.9% (7/143) and 4.2% (6/143) based on series testing using RBPT and c- ELISA, respectively. A seroprevalence of 21.7% (104/478) and 11.8% (33/278) based on RBPT were revealed in cattle and goats respectively. In contrast, c-ELISA revealed a seroprevalence of 21.3% (102/478) and 11.8% (33/278) in cattle and goats, respectively. Surprisingly, all the 86 serum samples from sheep tested on RBPT and further subjected to c-ELISA were found to be negative giving a seroprevalence of 0% (0/86) as shown in Table 1.

**Table 1:** Overall seroprevalence of brucellosis in human, cattle, goats and sheep based on RBPT and c-ELISA.

| Species | Number of sera tested | Seroprevalence |  |
| --- | --- | --- | --- |
|  |  | Total number of RBPT positive reactors n(%) | Total number of c-ELISA positive reactors n(%) |
| Human | 143 | 7 (4.8) | 6(4.2) |
| Cattle | 478 | 104 (21.7) | 102 (21.3) |
| Goats | 279 | 33 (11.8) | 33(11.8) |
| Sheep | 86 | 0 (0.0) | 0 (0.0) |

### Seroprevalence of brucellosis in cattle camps

The seroprevalence in the cattle camps of Terekeka County was 100% (13/13) compared to Juba County which was 50% (2/4) as shown in Table 2.The following species of livestock were sampled from the camps, cattle 56.7%, followed by goats at 33.1% and sheep 10.2%.Cattle were the dominant species in the camps of Terekeka County 89.5% (428/478) compared to Juba County 10.5% (50/478).

**Table 2:**
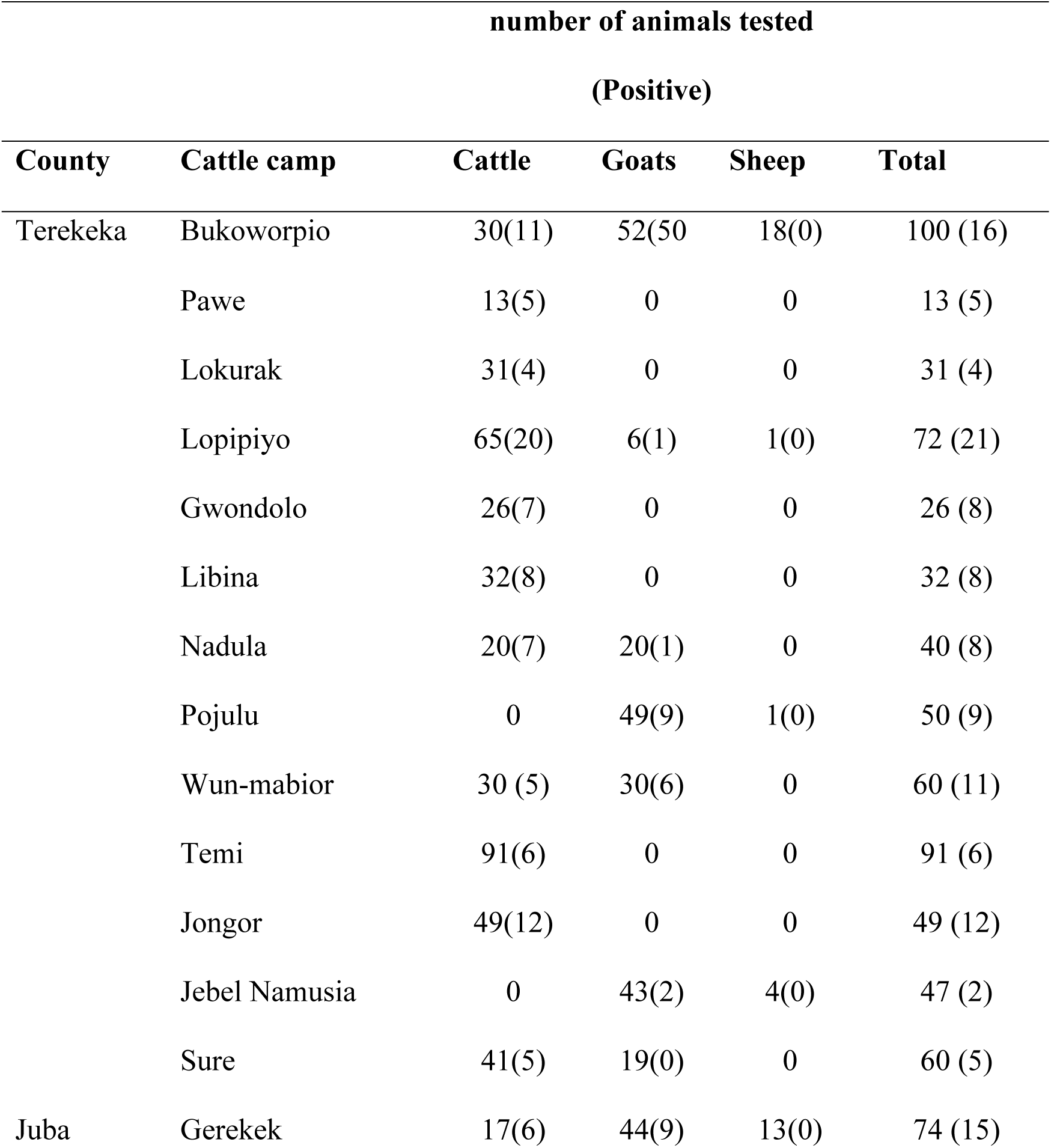
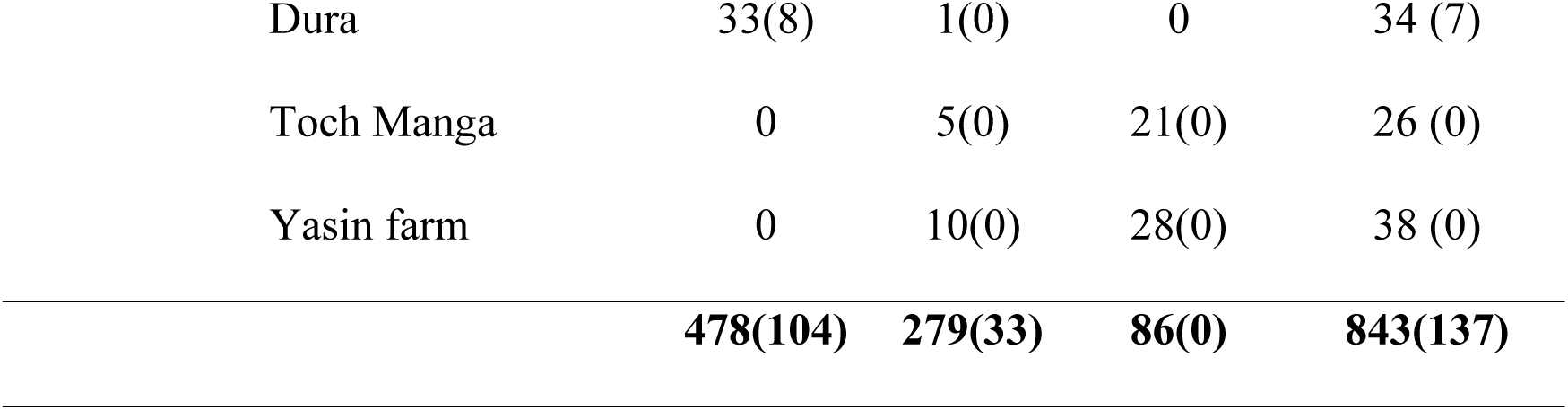
Seroprevalence of brucellosis in different Counties, Payams and cattle camps, CES, South Sudan number of animals tested.

### Risk factors associated with *Brucella* sero-positivity in humans, cattle and small ruminants in CES, South Sudan

#### Univariate logistic regression analysis

All the variables investigated in the univariate analysis of risk factors in cattle were significantly associated with *Brucella* seropositivity as shown in Table 3: sex (OR:4.5, 95% CI: 2.2 – 8.9, p:<0.001), age (OR:6.6, 95% CI: 2.3 – 19.1, p:<0.001), abortion history (OR:3.1, 95% CI: 1.8 –5.2, p:<0.001), retained placenta (OR:2.5, 95% CI: 1.4 – 4.4, p: 0.001), parity (OR:2.3, 95% CI: 1.142 – 4.739, p:0.02) and reproductive status (category “dry” OR:3.329, 95%CI: 1.598 – 6.934, p:0.001). The analysis shows a statistically significant association between sex and the occurrence of the condition at the univariate analysis. Females have a significantly higher likelihood of testing positive compared to males, as indicated by the low p-value (<0.001). There was also statistically significant association between age and the occurrence of the condition. Individuals over 5years old have the highest likelihood (OR: 6.6) of testing positive, followed by those aged 2-5 years old.

**Table 3:** Univariate analysis of potential risk factors associated with *Brucella* seropositivity in cattle in CES, South Sudan.

| Variable | Category | No.<br>sampled | No.<br>positive | OR | 95% CI | p-value |
| --- | --- | --- | --- | --- | --- | --- |
| Sex | Male | 132 | 10 |  |  |  |
|  | Female | 346 | 93 | 4.485 | 2.256 – 8.915 | < 0.001 |
|  | ≥ 1 year old | 75 | 4 |  |  |  |
| Age | 2 - 5 years old | 201 | 44 | 4.975 | 1.721 – 14.376 | 0.003 |
|  | >5 years old | 202 | 55 | 6.641 | 2.315 – 19.050 | < 0.001 |
| Abortion | No | 264 | 56 |  |  |  |
|  | Yes | 82 | 37 | 3.054 | 1.806 – 5.166 | <0.001 |
| Retained Placenta | No | 281 | 65 |  |  |  |
|  | Yes | 65 | 28 | 2.515 | 1.431 – 4.419 | 0.001 |
| Reproductive | Not | 91 | 17 | - | - | - |
| status | produced |  |  |  |  |  |
|  | Pregnant | 37 | 12 | 2.089 | 0.878 – 4.972 | 0.096 |
|  | Lactating | 158 | 38 | 1.378 | 0.726 – 2.617 | 0.326 |
|  | Dry | 60 | 26 | 3.329 | 1.598 – 6.934 | 0.001 |
|  | NA | 132 | 10 | - | - | - |
| Parity | Not | 90 | 17 | - | - | - |
|  | produced |  |  |  |  |  |
|  | Produced | 62 | 16 | 1.494 | 0.687 – 3.245 | 0.311 |
|  | once |  |  |  |  |  |
|  | Produced | 74 | 26 | 2.326 | 1.142 – 4.739 | 0.020 |
|  | twice |  |  |  |  |  |
|  | Produced | 120 | 34 | 1.698 | 0.877 – 3.286 | 0.116 |
|  | more than |  |  |  |  |  |
|  | twice |  |  |  |  |  |

Nevertheless, in the univariate analysis of risk factors associated with *Brucella* seropositivity in small ruminants, animal species (X^2^ = 11.183, p-value 0.001) and parity level (X^2^ = 10.394, P= 0.034) were found to be associated with *Brucella* seropositivity as showed in Table 4. The risk of occurrence of brucellosis in goats is higher compared to sheep as supported by the low p-value (<0.001).

**Table 4:**
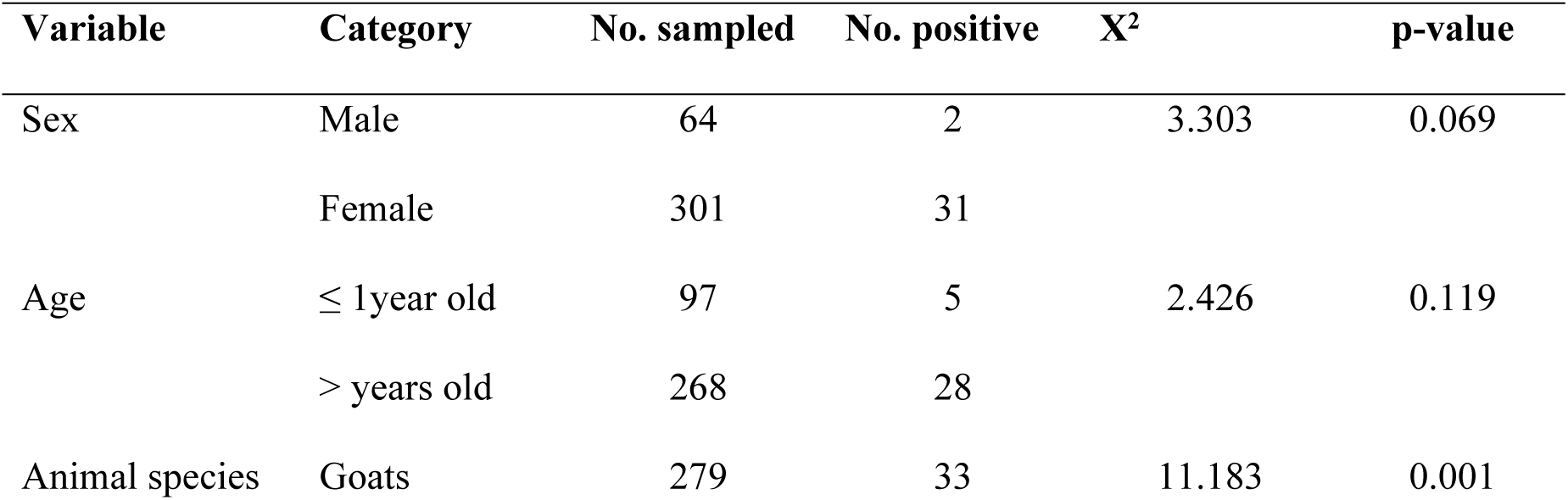
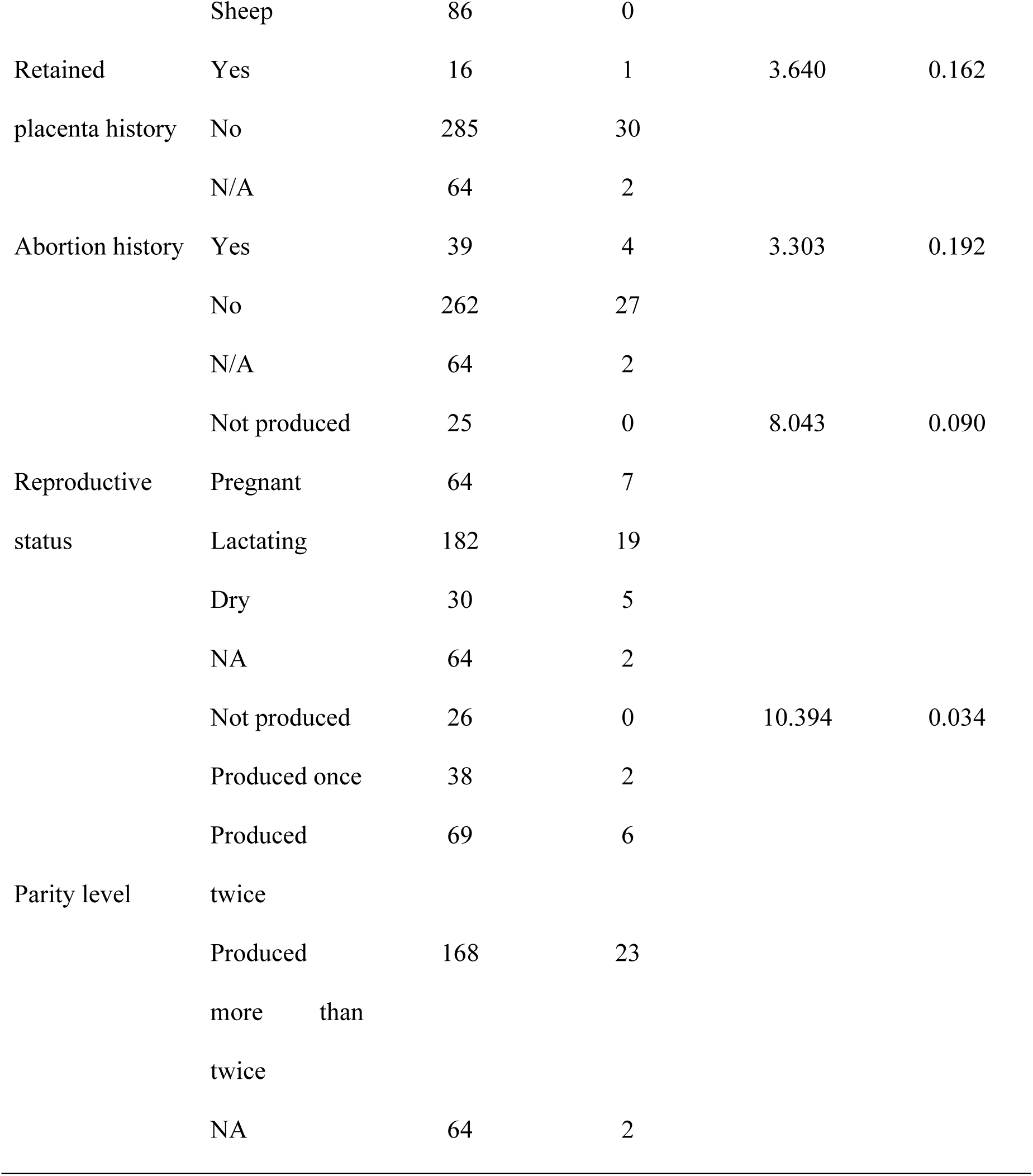
Univariable analysis of potential risk factors associated with *Brucella* seropositivity in small ruminants.

In humans, blowing air through cow’s vagina (OR: 1.4, 95%CI: 0.782 – 2.434, p: 0.035) was the only variable found to be significantly associated with *Brucella* seropositivity at the univariate analysis of risk factors.

Furthermore, variables with p-value≤2.5 were included in a multivariable regression analysis (Table 5).

**Table 5:**
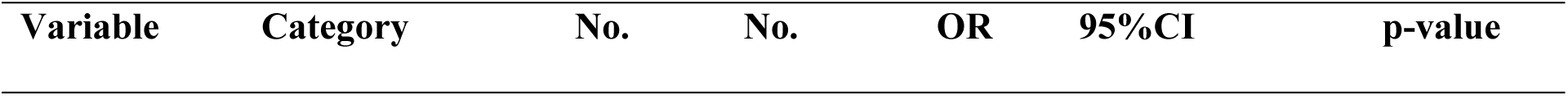
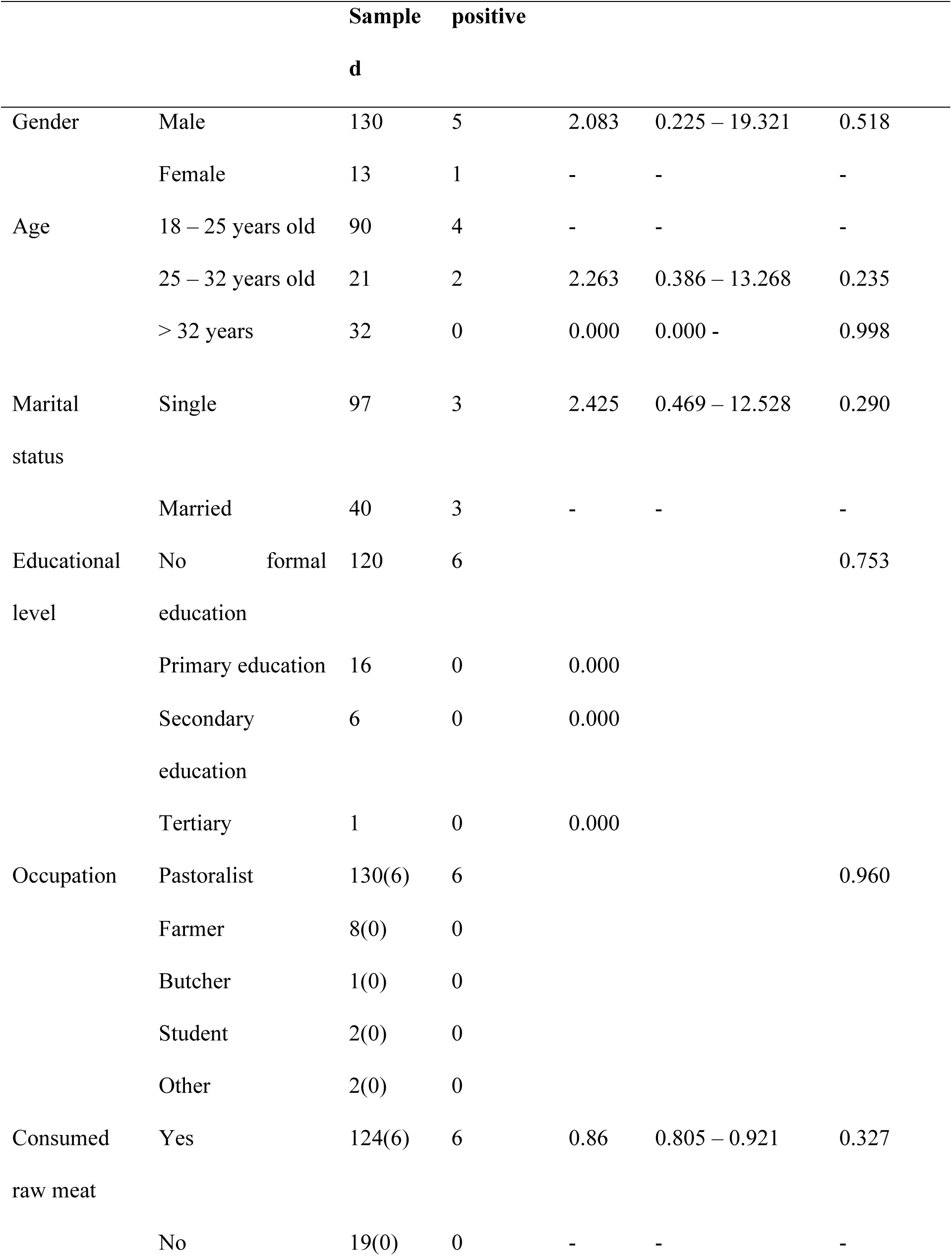
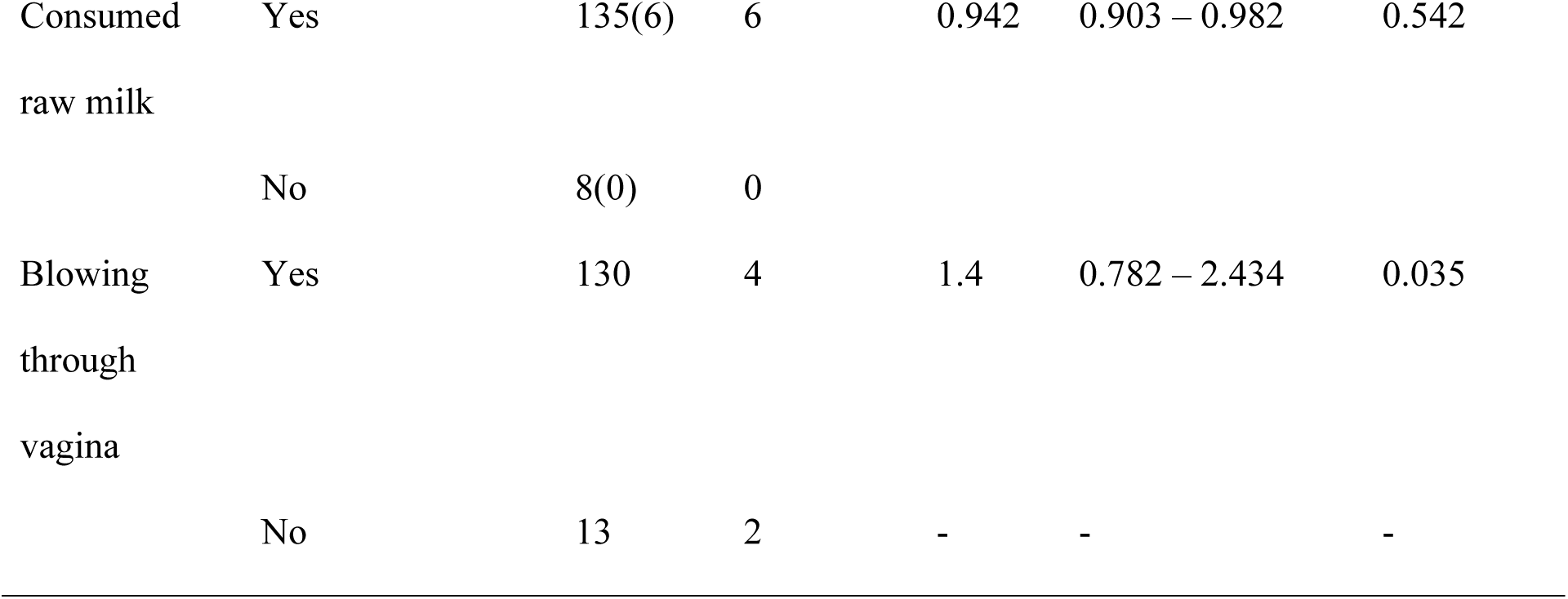
Univariate analysis of potential risk factors associated with *Brucella* seropositivity in humans.

### Multivariable logistic regression analysis

In cattle, all the six variables from the univariate analysis were included in the multivariable model. The multivariable logistic regression model identified sex, age, and abortion history as statistically significant factors of *Brucella* seropositivity in cattle. The odds of seropositivity were nearly threefold (OR: 2.8; 95% CI: 1.3 – 5.8, p: 0.006) higher in cows compared to bulls (male cattle). Older cattle over two years had higher odds of *Brucella* seropositivity than young animals (OR: 3.5, 95% CI: 1.2 – 10.3-, p: 0.025). Cows with a history of abortion had higher odds of *Brucella* seropositivity (OR: 2.8, 95% CI: 1.6 – 4.7, p: <0.001).

The Hosmer-Lemeshow goodness-of-fit test showed that the model fairly fitted the data (X^2^ = 10.281, p-value: 0.113).

However, in small ruminants, none of the variables was found to be statistically significant (P < 0.05) at the multivariate analysis with *Brucella* seropositivity.

**Table 5:**
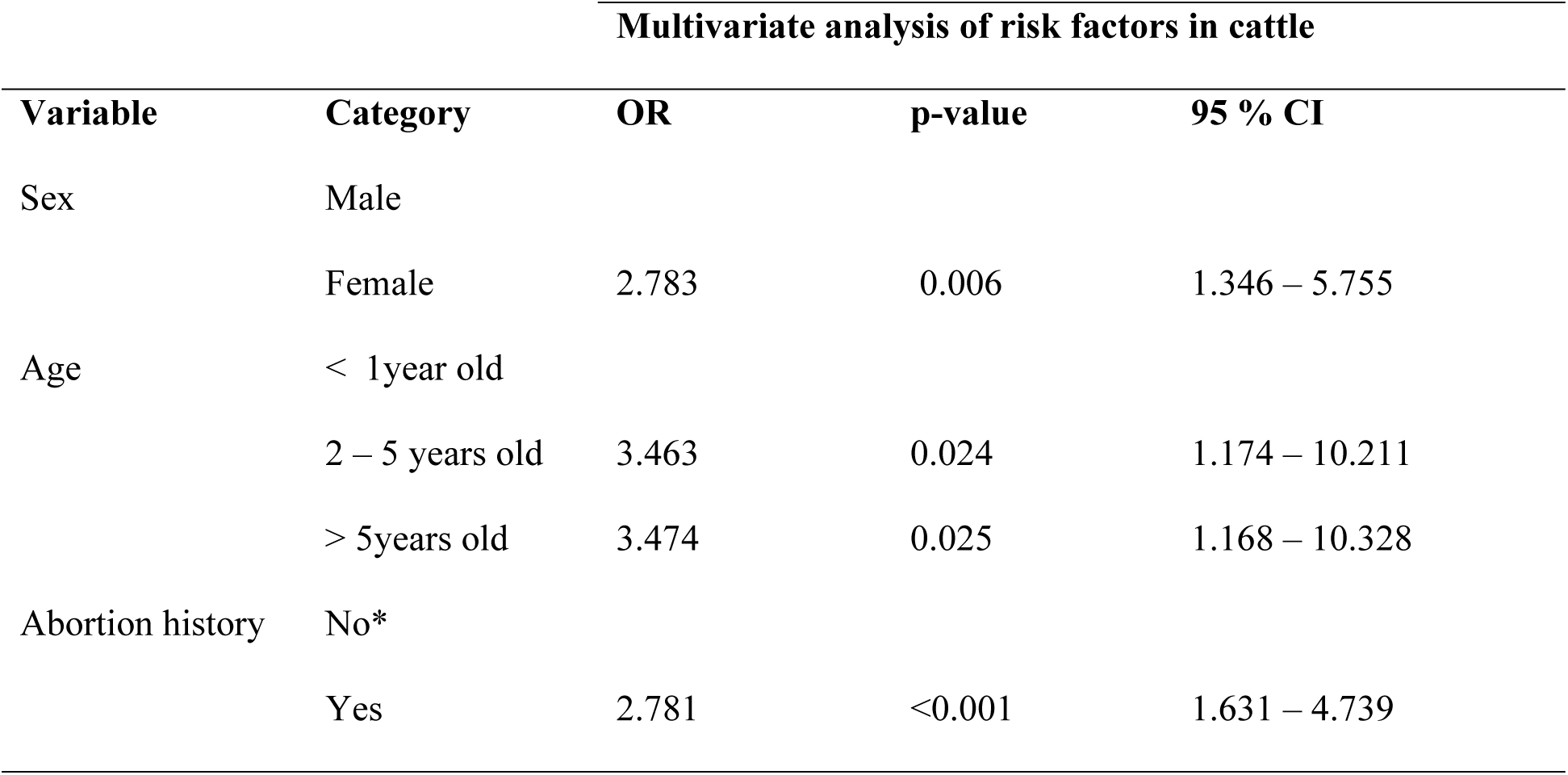
Multivariable logistic regression analysis of risk factors for *Brucella* seropositivity in cattle and small ruminants.

| Multivariate analysis of risk factors in cattle |  |  |  |  |
| --- | --- | --- | --- | --- |
| Variable | Category | OR | p-value | 95 % CI |
| Sex | Male |  |  |  |
|  | Female | 2.783 | 0.006 | 1.346 – 5.755 |
| Age | < 1year old |  |  |  |
|  | 2 – 5 years old | 3.463 | 0.024 | 1.174 – 10.211 |
|  | > 5years old | 3.474 | 0.025 | 1.168 – 10.328 |
| Abortion history | No* |  |  |  |
|  | Yes | 2.781 | <0.001 | 1.631 – 4.739 |

## DISCUSSION

This study has for the first time revealed seroprevalence of brucellosis in, goats, and the pastoral communities, and the absence of seropositive sheep in CES, South Sudan. The study revealed higher seroprevalence of brucellosis in cattle than in goats and identified the following risk factors; age, sex and previous history of abortion as significantly associated with *Brucella* seropositivity.

The current seroprevalence of brucellosis among the pastoral community was 4.1% (6/143) based on c-ELISA performed on RBPT seropositive sera. This seroprevalence is lower compared to the finding by [16] in Wau, Western Bahr el Ghazal state (WBeGS), who revealed a seroprevalence of 23.3% (97/416) based on c-ELISA. Additionally, a higher seroprevalence of 44% (56/126) based on serum agglutination test (SAT) was reported by [18] in Terekeka County, CES. These variations on the seroprevalence could be attributed to the nature and design of those previous studies. The seroprevalence reported in this study was community-based, participants were randomly selected, and the design was across-sectional compared to the previous studies, which were hospital-based and targeted suspected febrile patients. In Tanzania, a lower seroprevalence of 1.6 % in humans, has been reported by[24] and high prevalence of 7.7%reported by[25]. In Uganda on the other side,[26]reported a seroprevalence of 7.5% (15/200) among febrile out patients in Wakiso district, Central Uganda.

The variations on brucellosis seroprevalence could be attributed to spatial and temporal features and animal husbandry practices, pastoralists lifestyles, availability of veterinary services and control programs[9].

In South Sudan, livestock production systems are categorized as pastoral and agro-pastoral. A variety of livestock species including cattle, sheep and goats are reared collectively and kept in camping called ‘cattle camps’. The dominant species kept is cattle, followed by goats and to a lesser extent the sheep as this correlate with number sample collected in this study.

In cattle, the study revealed higher seroprevalence, 21.3% (102/478), compared to goats, 11.8% (33/279). This finding is in agreement with[24], those who reported a significantly higher prevalence in cattle than in goats in Tanzania.[25] had reported that cattle were more at risk of contracting *Brucella* infection than goats. Comparatively, the seroprevalence reported in cattle in this study is less compared to the 25.3% (86/340) reported by [19] and 29.3% (147/502) by [17]in South Sudan. However, another study reported a higher individual animal seroprevalence of 30.8% (88/285) and a lower herd prevalence of 77.7% compared to the findings of this study [12]. The high seroprevalence in cattle could be due to their dominance in the cattle camps in the study area. In South Sudan, cattle are kept for prestige, and the pastoralists rarely contemplate selling or culling them out. Hence, cattle harboring *Brucella* could have a chance of living longer than small ruminants in the cattle camps and would continue shedding infection given that the seroprevalence rises with age.

The study also revealed a seroprevalence of 11.8% (33/279) in goats. The seroprevalence was high in female goats 8.5% (31/365) compared to male goats 0.5% (2/365). Similarly, the prevalence of this study is in agreement with[27]who revealed higher prevalence of brucellosis in females 10.3% (31/301) compared to males 3.1% (2/64) in Arsi zone, Oromia, Ethiopia. [28]also reported a higher seroprevalence of brucellosis in female goats 1.4% (4/276) than males 0% (0/84) in Korahey zone, Somali regional state, eastern Ethiopia. The seroprevalence of goats in this study is also in agreement with a prevalence of 11.4% (35/307) reported by[15] on caprine in Khartoum State, Sudan. [29]in Borona pastoral areas in Southern Ethiopia reported higher prevalence 17.36% (137/789) of brucellosis in goats than the seroprevalence reported on this study.

Additionally,[10] reported a higher seroprevalence of 3.92% (13/332) in goats compared to 1.23% (1/81) in sheep in Karega District, Uganda.

The fact that none of the well-established risk factors for *B. melitensis* infection in goats were found associated with seropositivity in goats in our study suggests that not *B. melitensis* but most likely *B. abortus* spilling over from cattle could be the cause for seropositivity in goats. Indeed, although reports of *B. abortus* infection in small ruminants are scarce, such infections have been reported worldwide [30].

This study revealed a 0.0% seroprevalence of brucellosis in sheep (0/86). This is in line with [27] in Ethiopia who reported a 0.0% seroprevalence of brucellosis.

In West Africa, there is no report of *B. melitensis* infection in small ruminants. Seropositivity in small ruminants was documented in Nigeria to be associated with *B. abortus* infection that had spilled over from infected cattle [31]. In Latin America, sheep are not significantly infected with *B. melitensis* even when kept in close contact with goats[32]. Moreover, they do not easily become infected with *B. abortus* [33].This could be attributed to factors such as breed susceptibility, predominance species, husbandry practices and the self-limiting nature of the disease in sheep[34]. Reports from Egypt and Iran suggest that sheep are less susceptible to *B. abortus* infections than goats [30,35]

The fact that brucellosis seropositivity was not detected in sheep means that this species cannot be recognized as a source of human infection, which is an important epidemiological feature with implications in prospective One Health control measures. Moreover, it raises interesting questions regarding the etiology of brucellosis in South Sudan. In this perspective, a point of concern is the potential emergence of *Brucella* species infecting non-preferential hosts.

In the analysis of the risk factors, the study identified a significant association of *Brucella* seropositivity with sex, age, and abortion history in cattle. A Higher prevalence of brucellosis was identified in females cattle, 19.5 % (93/346) compared to males, 2.1% (10/132), and this difference was statistically significant (OR = 2.783, P-value < 0.006). This finding is in agreement with [13]who reported a significant association of *Brucella* seropositivity with sex on which female animals had higher level of exposure compared to males. Other researchers have reported similar findings of significant association of *Brucella* seropositivity in female animals[24,36].

The current study also revealed that cows with a prior history of abortion had higher odds of *Brucella* seropositivity (OR: 2.8, 95% CI: 1.6 – 4.7, p: <0.001). Our finding is in agreement with a previous study conducted in South Sudan [17]as well as with the findings from multiple studies[37–40] which reported an association of *Brucella* seropositivity with abortion.

This could be due to repeated exposure to *Brucella* spp. as female animals stay for longer periods in herds than males. Furthermore, the female reproductive tract provides a potential reservoir for the organism to propagate due to the presence of erythritol sugar which stimulates the growth of *Brucella* organism. The study revealed that, older cattle over two years of age (OR: 3.5, 95% CI: 1.2 – 10.3-, p: 0.025) had higher odds of *Brucella* seropositivity than younger ones. This finding is in agreement with several studies [37,38] that also identified age as a risk factor for *Brucella* seropositivity in cattle. In contrast to the current’s study finding, another study had revealed a higher odds of *Brucella* infection in young compared to adults [41].

The fact that older cattle showed higher seropositivity to *Brucella* infection than the young ones could be attributed to continued exposure to pathogen, especially in the cattle camps where cattle are kept over long periods. The seroprevalence of brucellosis in the herds within cattle camps of Terekeka County was 100% compared to Juba County which was 50%. This finding is in agreement with [17] who reported herd seroprevalence based on c-ELISA at 61.4% and 90.0% for peri-urban Juba town and rural Terekeka County cattle herds, respectively. Our results suggest that cattle are a reservoir of brucellosis in livestock, because of the highest seroprevalence found in this species, most likely due to *B. abortus*, its preferential host. The lower seroprevalence in goats suggests that *B abortus* may have spilled over from cattle to goats. The absence of seropositivity in sheep suggest that *B. melitensis* not endemic in this species and that *B. abortus* has not yet spilled over to the sheep due to the husbandry systems, with spatial and temporal segregation mainly between cattle and sheep.

## CONCLUSION

This study report for the first time seroprevalence of brucellosis in goats in South Sudan where it’s prevalence in livestock and pastoral community revealed its endemicity.

Female cattle have a higher risk of infection compared to males. Previous history of abortion and older cattle were significantly associated with *Brucella* seropositivity. Based on our findings, we recommend that control measures should be directed to cattle to reduce production losses and possible spillover to goats and to prevent human contamination. Moreover, strategies for nationwide awareness campaigns and implementing One Health approach are needed to mitigate brucellosis in South Sudan effectively.

Efforts should be put on the isolation of *Brucella* spp. from cattle and goats to document that *B. abortus* has spilled over from its cattle reservoir to goats and to prevent its further spill over to sheep.

Additionally, control measures should first be directed to cattle to reduce production losses and possible spillover to goats and to prevent human contamination from a One Health perspective.

## ETHICAL CLEARANCE

The study protocol was approved by the Institutional Review Board of Sokoine University of Agriculture under reference number (DPRTC/R/186/16) and the National Ministry of Health Research Ethics Review Board (RERB-P No: 13/14/02/2023), South Sudan.

Additionally, permissions for data collection were obtained from the State Ministry of Animal Resources, Fisheries and Tourism and the Ministry of Health, CES, South Sudan.

Moreover, Export and Import permits for shipment of the biological samples were obtained from the National Ministry of Livestock and Fisheries, South Sudan, and the Ministry of Livestock and Fisheries, United Republic of Tanzania.

Informed consent was obtained from each study participant who agreed to participate in the study prior to data collection.

## Data Availability

All datasets used or analyzed during the current study are available from the corresponding author upon request.

## ACKNOWLEDGMENTS

The authors are grateful to acknowledge the State Ministries of Health and that of Animal Resources, Fisheries and Tourism in CES, Juba for granting permission for data collection. We would like to appreciate the National Ministries of Livestock and Fisheries and of Trade and Industries in South Sudan for issuing an Export Permit. The Ministry of Livestock and Fisheries in Tanzania is greatly appreciated for issuing the Import permit of the biological samples. We are indebted to Sokoine University of Agriculture for availing laboratory facility. Additionally, we would like to express our appreciation to all field and laboratory technicians for technical assistance. Many thanks, to Dr. Lazarus Lugoi, University of Juba for producing the map of the study area. We acknowledge the cattle owners in the cattle camps for their effective participation and amicable cooperation during data collection.

